# Direct diagnostic testing of SARS-CoV-2 without the need for prior RNA extraction

**DOI:** 10.1101/2020.05.28.20115220

**Authors:** Shan Wei, Esther Kohl, Alexandre Djandji, Stephanie Morgan, Susan Whittier, Mahesh Mansukhani, Eldad Hod, Mary D’Alton, Yousin Suh, Zev Williams

## Abstract

The COVID-19 pandemic has resulted in an urgent global need for rapid, point-of-care diagnostic testing. Existing methods for nucleic acid amplification testing (NAAT) require an RNA extraction step prior to amplification of the viral RNA. This step necessitates the use of a centralized laboratory or complex and costly proprietary cartridges and equipment, and thereby prevents low-cost, scalable, point-of-care testing. We report the development of a highly sensitive and robust, easy-to-implement, SARS-CoV-2 test that utilizes isothermal amplification and can be run directly on viral transport media following a nasopharyngeal swab without the need for prior RNA extraction. Our assay provides visual results in 30 min with 85% sensitivity, 100% specificity, and a limit of detection (LoD) of 2.5 copies/μl, and can be run using a simple heat block.

## Introduction

The Coronavirus disease 2019 (COVID-19) pandemic caused by SARS-coronavirus 2 (SARS-CoV-2) has created a global health emergency with more than five million confirmed cases and more than 300,000 deaths as of May 25, 2020^1^. Widespread molecular diagnostic testing for the virus is crucial for a managed response to the ongoing SARS-Cov-2 pandemic, including prompt diagnosis, contact-tracing, quarantine, and treatment^2^, especially given the apparently high rate of asymptomatic viral shedding^3^. The primary method of testing for active disease has been various forms of nucleic acid amplification tests (NAAT). Existing detection methods based on RT-PCR, isothermal amplification, or Crispr-Cas12, all require an initial RNA extraction step prior to nucleic acid amplification^4–7^. As a result, samples must either be transported to a centralized high-complexity laboratory or processed at the point-of-care using systems that rely on specialized, proprietary instruments and consumables, limiting the capacity to scale testing for widespread use both in the U.S. and globally^8^. There is, therefore, a critical need for a single-step SARS-CoV-2 diagnostic test that does not require prior RNA extraction and that can be performed directly on clinical samples at the point-of-care with prompt delivery of results and without the need for costly and scarce proprietary equipment and reagents.

Loop-mediated isothermal amplification (LAMP) is a targeted nucleic acid amplification method that utilizes a combination of primer sets and a DNA polymerase with high strand displacement activity to specifically replicate a region of DNA^9^. LAMP has several distinct advantages for point-of-care testing of clinical samples for SARS-CoV-2. While traditional PCR requires a costly and complex thermocycler, the entire LAMP amplification reaction is performed at a single temperature, and thus requires only a heat block or water bath. The polymerase enzyme used in LAMP (*Bst*) is more robust than that used in traditional PCR, and the product can be visualized with the unaided eye in real time using colorimetry, turbidity, or fluorescence with the aid of a fluorescent light source. In order to allow for the detection of RNA targets, such as SARS-CoV-2, a reverse transcription (RT) step (RT-LAMP) can be implemented in the same master mix at the start of the reaction.

Here we report the development and initial validation of a one-step SARS-CoV-2 detection test that can be run directly on clinical nasopharyngeal swab samples in transport media without the need for prior RNA extraction, and thus can be readily implemented for low-cost, point-of-care, SARS-CoV-2 testing.

## Methods and Materials

### RT-LAMP Assay Reagents preparation

Primer sets, buffers, and incubation methods were systematically tested to develop the optimized method used herein that would be sufficiently sensitive and robust to enable direct detection of viral RNA from clinical samples. PrimerExplorer (https://primerexplorer.jp/e/) and the SARS-Cov-2 reference genome NC_045512v2 were used to design the LAMP primers. A 25-fold primer mix of LAMP primers (CUFC1-FIP, CUFC1-BIP, CUFC1-LF, CUFC-LB, CUFC1-F3, CUFC1-B3; Table 1) was prepared by assembling 40μM FIP and BIP, 10μM CUFC1-LF and CUFC1-LB, and 5μM CUFC1-F3 and CUFC1-B3 primers in nuclease-free water (Ambion, AM9937). A 2X colorimetric RT-LAMP master mix was prepared by adding 3.5μL 100mM dUTP, 0.5μL UDG, and 0.25μL 5mM SYTO 9 (Invitrogen, S34854) to 1,250μL WarmStart® Colorimetric LAMP 2X Master Mix (DNA & RNA) (NEB, M1800L). The reaction mix for one 250μL reaction was prepared by mixing 125μL 2X colorimetric RT-LAMP master mix, 10μL 25-fold LAMP primer mix, and 95μL nuclease-free water. These values can be scaled up according to the actual number of samples; two 250μL reactions were used to test one sample. The lysis buffer consisted of 0.1-fold TE buffer pH 8.0 with 0.1% tween-20, 1% volume (e.g., 1μL enzyme added to 100μL buffer) Thermolabile Proteinase K (NEB, P8111S), 2% volume ezDNase (Invitrogen, 11766051), and 0.3ng/μL human genomic DNA from a normal male. For one reaction, 460μL of reaction mix and 20μL of lysis buffer were preloaded in a clean 1.5mL LoBind microcentrifuge tube (Eppendorf, 022431021) and kept on ice until use.

**Table 1.**
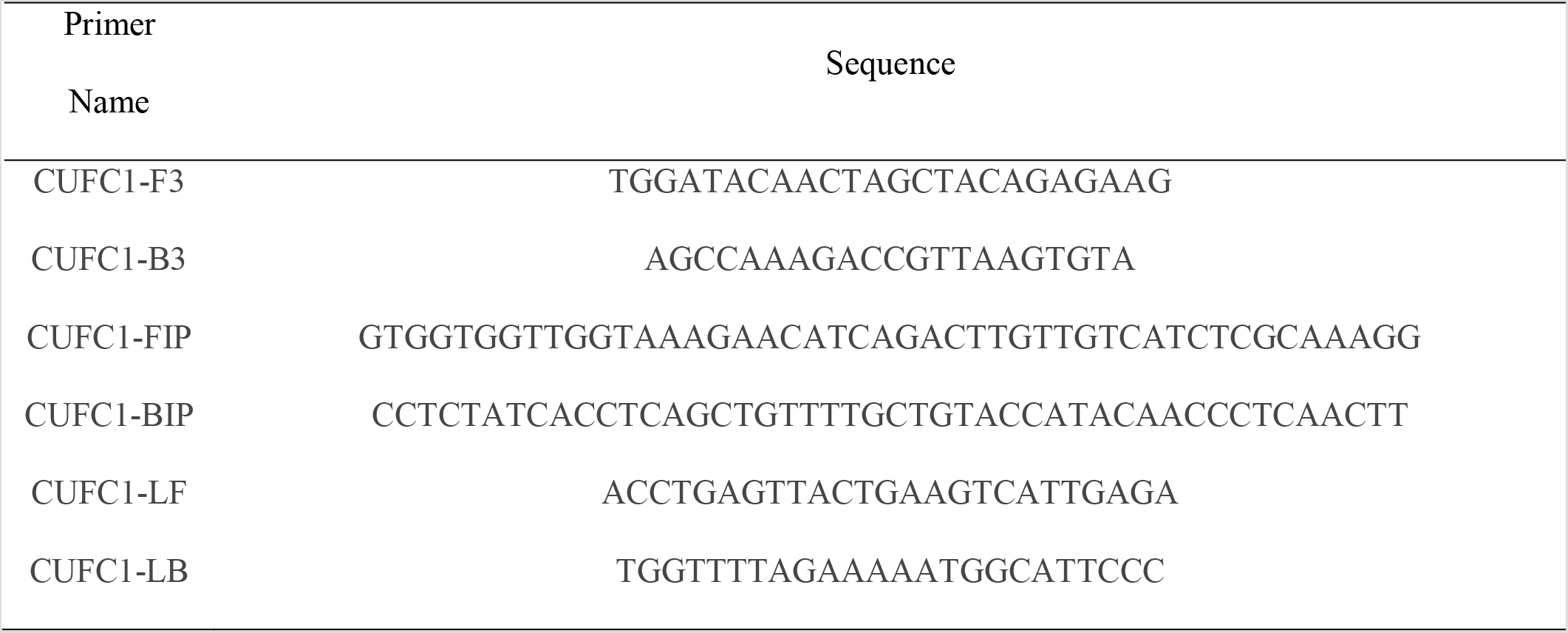
Sequence information of LAMP primers for SARS-Cov-2 detection Primer

### Initial testing of contrived and selected samples

The limit of detection (LoD) was determined by testing serial dilutions of the SARS-Cov-2 RNA standard (Exact Diagnostics, COV019) spike-in with viral transport medium (CDC SOP#: DSR-052–02), instead of clinical samples, following the optimized protocol detailed above.

To determine the LoD for clinical samples, a set of 20 positive clinical samples, selected to represent the range of Ct values detected using a Roche COBAS 6800 system for SARS-Cov-2, and 10 negative samples were subjected to the optimized LAMP protocol.

### Testing of Clinical Samples

Twenty remnant clinical samples consisting of viral transport media inoculated with a nasopharyngeal swab sample obtained as part of routine clinical testing were chosen at random. From each clinical specimen, 20 μL was placed directly into a 1.5mL LoBind microcentrifuge tube (Eppendorf, 022431021) containing the reaction mix (460μL) and lysis buffer (20μL). The solution was mixed using a sterile disposal transfer pipette (Fisherbrand, 13–711–20) by gentle pipetting 12 times. Using the same sterile disposable transfer pipette, 250uL of the 500 uL solution was placed into a new 1.5mL LoBind microcentrifuge tube. Both tubes with ∼250μL each were placed in a 63.0°C dry bath (Fisherbrand, 14–955–219) and incubated for 30 min. The tubes were then placed on ice for 1 min to pause the reaction and the colorimetric results were read (red=negative,yellow=positive).

## Results

We designed and optimized primers and reaction conditions for a high performance, direct, rapid colorimetric RT-LAMP test for SARS-CoV-2 (Figure 1). The final primer set targets the middle of *ORF1ab*, the largest SARS-Cov-2 gene (Figure 1A), and has a relatively low GC%. The optimal reaction temperature was determined experimentally to be 63°C. The workflow enables direct testing of clinical samples without the need for RNA isolation or cell lysis (Figure 1B)^7,10^. The set-up requires only a pipette and tips, a transfer pipette, a mini heat block, and a box of ice (Figure 1C); no special equipment or devices are needed. We used a colorimetric output for simple interpretation of results without the need for extra equipment. RT-LAMP amplification of the targeted DNA results in decreased pH, causing a pH sensitive dye in the reaction mixture to change color from red to yellow in positive tests while remaining red in negative tests (Figure 1D). For samples with a low copy number of virus, a positive signal was displayed only in one of the two tubes, in which case we interpreted the results as positive.

**Figure 1.**
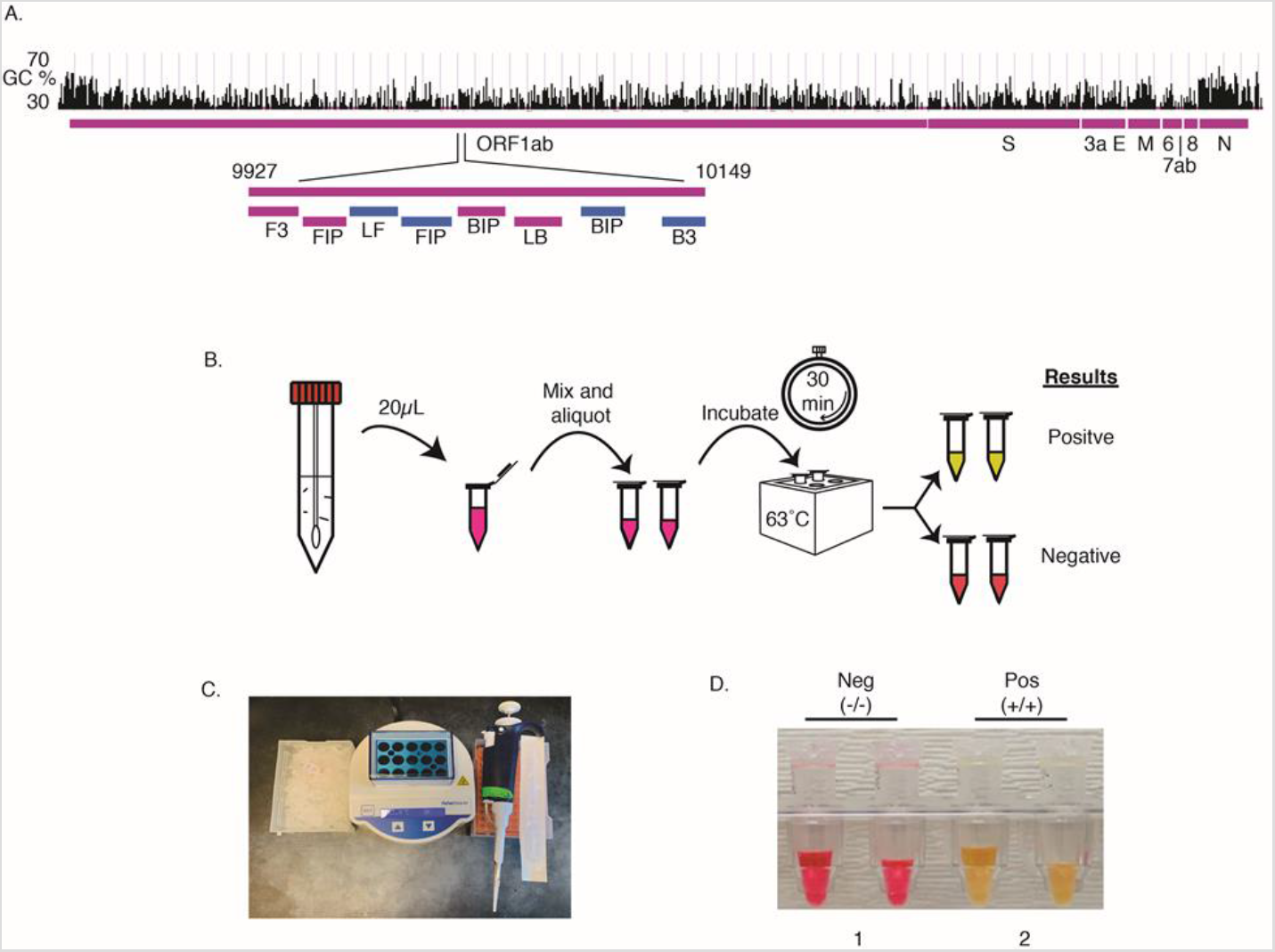
An RT-LAMP assay for rapid and direct SARS-Cov-2 testing of clinical samples at the point-of-care. A) Primer design. A set of 6 LAMP primers targeting the middle of the Orf1ab gene. Sequences and primers matching to the + strand of virus genome are in pink, while those matching to the – strand are shown in blue. B) Workflow of direct RT-LAMP assay. 20μL of raw clinical samples from nasal swab in viral transport media (VTM) was added to a 480μL reaction solution consisting of reaction master mix and lysis buffer. The resulting solution was mixed and 250 μL were aliquoted with a transfer pipette and then incubated on a heatblock for 30 min. The reaction was stopped by placing the samples on ice and results were interpreted by color due to a pH sensitive dye in the master mix (yellow=positive; red=negative). C). Setup of the RT-LAMP assay. D). An example of colorimetric results for negative and positive clinical samples (Ct 31.58 for target 1, 32.87 for target 2).

Viral transport medium contains inhibitors that reduce amplification sensitivity; in the RTLAMP reaction a 30 to 100-fold reduction in sensitivity was observed in comparison to buffers such as HBSS^7^. Nonetheless, samples collected as part of clinical care that had been placed in viral transport media were used to (a) keep the existing workflow as consistent as possible and(b) have a single nasopharyngeal swab sample tested in parallel using our test and the Roche COBAS system. To determine the LoD of our assay, we spiked-in viral RNA standards into viral transport media (Figure 2A). Serial dilution experiments, conducted in quadruplicate, consistently showed positive results down to 2.5 copies/μL. Results with copy numbers below 2.5 copies/μL were inconsistent, and thus 2.5 copies/μL was determined to be the LoD (Figure 2A).

**Figure 2.**
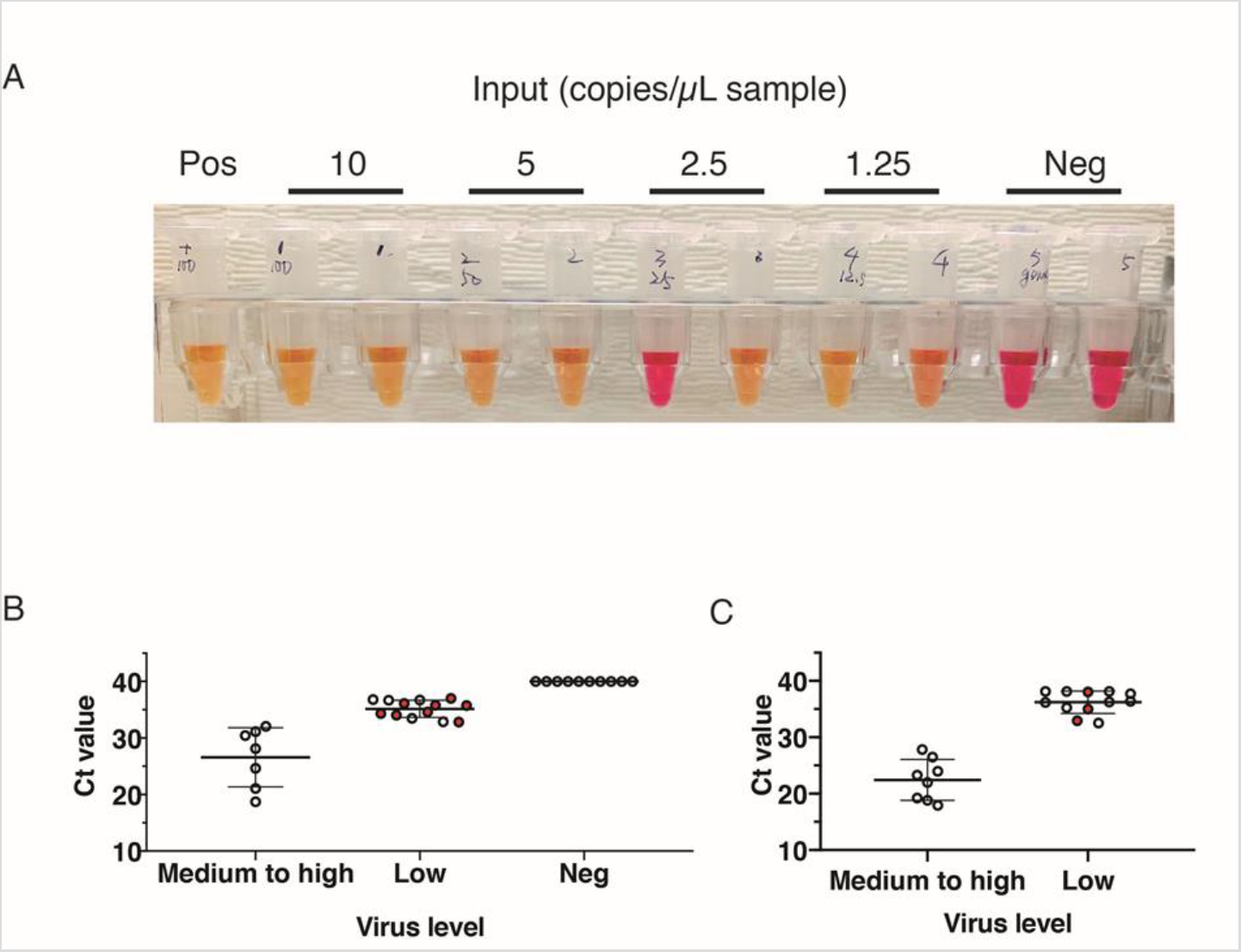
The performance of RT-LAMP testing for SARS-Cov-2. A) Estimation of limit of detection using VTM and RNA standard spike-ins. (Colorimetric results: red=negative; yellow=positive) B) Estimation of limit of detection using clinical samples. Each dot represents one sample, and samples with discordant testing results labeled in red. Error bars indicate Mean ± SD. C) Testing on random clinical samples. Each dot represents one sample, and samples with discordant testing results labeled in red. Error bars indicate Mean ± SD.

To compare the Ct values of clinical samples tested on the Roche COBAS system with our RT-LAMP results, positive clinical samples from routine clinical testing for COVID-19 were selected to represent the broad range of Ct value and were tested using our LAMP assay (Figure 2B). These samples showed Ct values ranging from 18.52 to 34.42 for RT–PCR Target 1, and 18.69 to 36.61 for RT–PCR Target 2 on the Roche COBAS 6800 system. Ten negative clinical samples were also tested. Samples with Ct value of ≤30 for target 1 and ≤31 for target 2, corresponding to ∼ 0.168 TCID_50_/mL (median tissue culture infectious dose), had stable performance and high accuracy in testing results (Figure 2B). Hence the LoD of clinical samples using our RT-LAMP assay is Ct ≤ 30 for RT-PCR target 1 and ≤ 31 for RT-PCR target 2. Five out of 13 samples with high Ct values tested positive, and not all of the false negative samples were necessarily the ones with highest Ct value, indicating that the presence of detectable virus was not within the linear range due to low copy numbers. All 10 of the clinically negative samples also tested negative by RT-LAMP, indicating that the assay has a specificity of 100% (Figure 2B, Table 2).

**Table 2.**
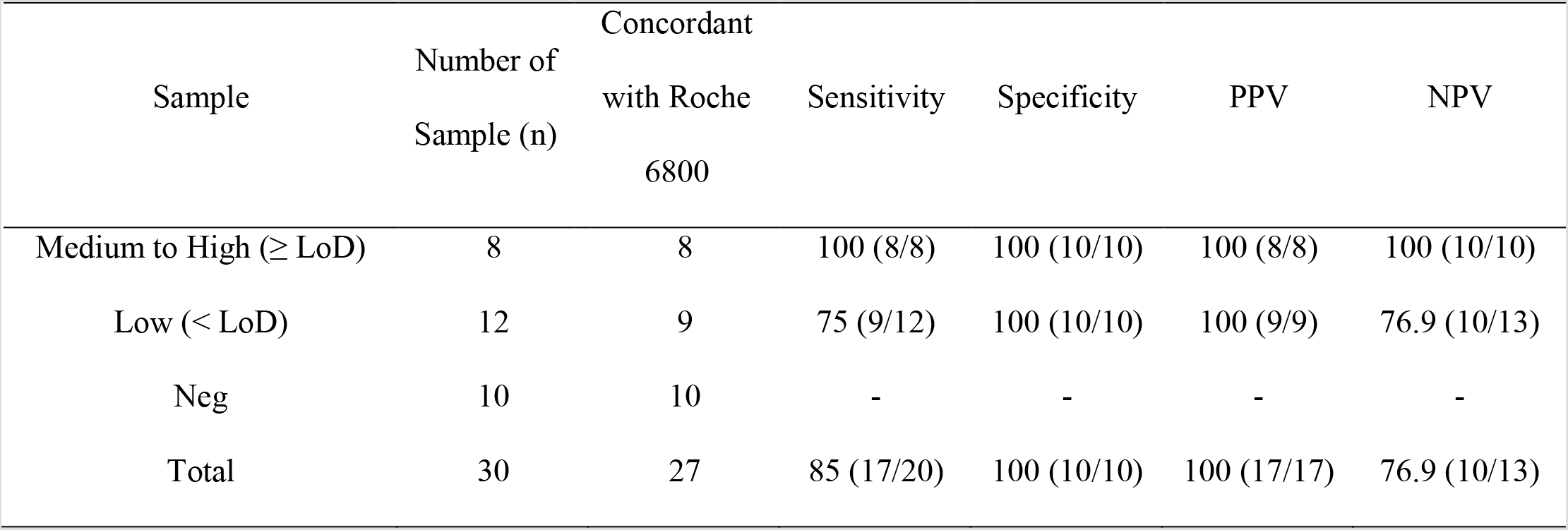
Summary of direct RT-LAMP testing on clinical samples.

To estimate the detection power of our RT-LAMP test on actual clinical specimens, 20 clinical samples tested positive and 10 clinical samples tested negative by standard test were randomly selected and subjected to the RT-LAMP assay. Samples had a Ct value ranging from 17.46 to 35.71 for Target 1, and 17.94 to 38.12 for Target 2. Eight samples were within, while 12 samples were below, the predicted LoD of the rapid testing method (Figure 2C). Eight out of 8 samples within the LoD tested positive and were concordant with clinical testing results. Nine out of 12 samples below the LoD tested positive. For samples below the LoD, our results indicate that RT-LAMP has 75% (9/12) sensitivity, 100% specificity (10/10), 100% (9/9) PPV, 76.9% (10/13) NPV, and 86.4% (19/22) accuracy (Table 2). In total, the rapid test generated an 85% (17/20) sensitivity, 100% (10/10) specificity, 100% PPV (17/17), 76.9% (10/13) NPV, and 90% (27/30) accuracy for randomly selected clinical samples (Table 2).

## Discussion

Here we developed a robust and highly sensitive method based on RT-LAMP for direct detection of SARS-CoV-2 in viral transport media in 30 min with a LoD as low as 2.5 copies/μl. A unique feature of our assay is that it does not require RNA isolation and/or cell lysis, and therefore can be applied directly to clinical samples, unlike other recent reports using RT-LAMP, RT-PCR or CRISPR-Cas12 for SARS-CoV-2 testing^10,11^. The ability to test at the point-of-care and return results in 30 min without the need for RNA extraction/purification or specialized equipment has practical advantages for on-site screening and detection of those with a higher viral load. In particular, this RT-LAMP test may be useful for quick primary screening of patients in the early stages of infection, when they are normally reported to have a viral load of 10^4^ to 10^7^ copies/mL^12^, who may be asymptomatic and spreading the virus. Thus, this method would also lend itself to widespread testing and testing in resource-poor settings.

Our assay was highly specific with no false positive results observed, but was not as sensitive as the Roche COBAS 6800 system. However, our assay had a higher positive agreement than the Abbott ID Now in samples with medium to low Ct values (100% vs 73.9%), and especially in samples with high Ct values (76.9% vs. 34.3%)^13^. In contrast to the Abbott ID Now, our approach does not require specialized equipment and cartridges and hence may be more readily scaled and used globally without the need to manufacture and ship the specific hardware and can use reagents available from multiple manufacturers located internationally. Further optimization in primer design, adjustments to buffers, multiplex primers targeting multiple regions of the virus, and other adjustments, improving the sensitivity of the assay as well as validation would be required to improve the assay. Viral transport media contains inhibitors that reduced the sensitivity of our assay by ∼30x, but we still chose to use nasopharyngeal swab samples placed in viral transport media for this study so that the testing could be readily incorporated into the existing testing workflow and so that a single sample could be processed using both our method and the Roche COBAS 6800 system for validation. For use as a point-of-care test where transport is not needed, using a more basic buffer such as HBSS or direct placement of the swab into the master mix reaction buffer could make this processes simpler, faster and more sensitive. We utilized a pH sensitive colorimetric indicator to provide a very simple visual read-out. However, our method would also be compatible with other outputs such as turbidity, fluorescence or recently developed CRISPR-Cas12-based indicators^7^.

To successfully manage the COVID-19 pandemic, cost-effective, efficient, and frequent screening is necessary worldwide. The speed, ease-of-use, scalability, and use of widely-sourced reagents make our high performance RT-LAMP method well-suited for initial screening at the point of care and at population levels.

## Data Availability

No sequencing data is generated in this study. Data referred to is available upon request.

## Acknowledgement

We thank Drs. Kevin Roth and Steven Spitalnik, of the Department of Pathology and Cell Biology and members of the Williams Laboratory, Columbia University Fertility Center, Suh Laboratory, Clinical Microbiology Laboratory, Columbia University Laboratory of Personalized Genomic Medicine at Columbia University Medical Center and New York Presbyterian Hospital for their helpful inputs and support for this study. This study is supported by NIH grants R01HD100013 (Z.W.), R01HD086327 (Z.W.), 1RF1AG057341 (Y.S), and R01AG057433 (Y.S).

## References

1. World Health Organization. Coronavirus disease (COVID-19) Situation Report-103,<https://www.who.int/docs/default-source/coronaviruse/situation-reports/20200502-covid-19-sitrep-103.pdf?sfvrsn=d95e76d8_6> (2020).

2. Salathe, M. et al. COVID-19 epidemic in Switzerland: on the importance of testing, contact tracing and isolation. Swiss Med Wkly 150, w20225, doi:10.4414/smw.2020.20225 (2020).

3. He, X. et al. Temporal dynamics in viral shedding and transmissibility of COVID-19. Nat Med, doi:10.1038/s41591-020-0869-5 (2020).

4. Sheridan, C. in Nat Biotechnol (2020).

5. Yan, C. et al. Rapid and visual detection of 2019 novel coronavirus (SARS-CoV-2) by a reverse transcription loop-mediated isothermal amplification assay. Clin Microbiol Infect, doi:10.1016/j.cmi.2020.04.001 (2020).

6. Wang, J. et al. A multiple center clinical evaluation of an ultra-fast single-tube assay for SARS-CoV-2 RNA. Clin Microbiol Infect, doi:10.1016/j.cmi.2020.05.007 (2020).

7. Broughton, J. P. et al. CRISPR–Cas12-based detection of SARS-CoV-2. Nature Biotechnology, 1–5, doi:10.1038/s41587-020-0513-4 (2020).

8. Cheng, M. P. et al. Diagnostic Testing for Severe Acute Respiratory Syndrome-Related Coronavirus-2: A Narrative Review. Ann Intern Med, doi:10.7326/m20-1301 (2020).

9. Notomi, T. Loop-mediated isothermal amplification of DNA. Nucleic Acids Research 28, 63e–63, doi:10.1093/nar/28.12.e63 (2000).

10. Zhang, Y. et al. Rapid Molecular Detection of SARS-CoV-2 (COVID-19) Virus RNA Using Colorimetric LAMP. medRxiv, 2020.2002.2026.20028373–20022020.20028302.20028326.20028373, doi:10.1101/2020.02.26.20028373 (2020).

11. Yang, W. et al. Rapid Detection of SARS-CoV-2 Using Reverse transcription RT-LAMP method. medRxiv, 2020.2003.2002.20030130–20032020.20030103.20030102.20030130, doi:10.1101/2020.03.02.20030130 (2020).

12. Wyllie, A. L. et al. Saliva is more sensitive for SARS-CoV-2 detection in COVID-19 patients than nasopharyngeal swabs. medRxiv, 2020.2004.2016.20067835–20062020.20067804.20067816.20067835, doi:10.1101/2020.04.16.20067835 (2020).

13. Smithgall, M. C., Scherberkova, I., Whittier, S. & Green, D. A. Comparison of Cepheid Xpert Xpress and Abbott ID Now to Roche cobas for the Rapid Detection of SARS-CoV-2 bioRxiv, doi:https://doi.org/10.1101/2020.04.22.055327 (2020).

